# A familial modeling framework for advancing precision medicine in neuropsychiatric disorders: A study in children with RASopathies

**DOI:** 10.1101/2024.02.06.24302411

**Authors:** Jennifer L. Bruno, Jacob Joseph Merrin, Hadi Hosseini, Tamar Green

**Author notes:** Address correspondence to: Jennifer Bruno, Department of Psychiatry and Behavioral Sciences, Stanford University School of Medicine, 401 Quarry Rd., Stanford, CA 94305-5795, USA. **Study Type**: Cross Sectional. **Data Sharing statement:** The final dataset will be stripped of all identifiers and made available to qualified investigators upon request.

## Abstract

**Objective:** Despite the significant and growing burden of childhood psychiatric disorders, treatment is hindered by lack of evidence-based precision approaches. We utilized parent cognitive and behavioral traits in a predictive framework to provide a more individualized estimate of expected child neuropsychiatric and neuroanatomical outcomes relative to traditional case-control studies. We examined children with Noonan Syndrome, a neurogenetic syndrome affecting the Ras/mitogen-activated protein kinase (Ras/MAPK), as a model for developing precision medicine approaches in childhood neuropsychiatric disorders.

**Methods:** Participants included 53 families of children with Noonan syndrome (age 4-12.9 years, mean = 8.48, SD = 2.12, 34 female). This cross-sectional study utilized univariate regression to examine the association between non carrier parent traits (cognition and behavior) and corresponding child traits. We also used multivariate machine learning to examine the correspondence between parent cognition and child multivariate neuroanatomical outcomes. Main outcome measures included child and parent cognition, anxiety, depression, attention-deficit hyperactivity (ADHD) and somatic symptoms. We also included child neuroanatomy measured via structural MRI.

**Results:** Parent cognition (especially visuospatial/motor abilities), depression, anxiety and ADHD symptoms were significantly associated with child outcomes in these domains. Parent cognition was also significantly associated with child neuroanatomical variability. Several temporal, parietal and subcortical regions that were weighted most strongly in the multivariate model were previously identified as morphologically different when children with NS were compared to typically developing children. In contrast, temporal regions, and the amygdala, which were also weighted strongly in the model, were not identified in previous work but were correlated with parent cognition in post-hoc analysis suggesting a larger familial effect on these regions.

**Conclusions:** Utilizing parent traits in a predictive framework affords control for familial factors and thus provides a more individualized estimate of expected child cognitive, behavioral, and neuroanatomical outcomes. Understanding how parent traits influence neuroanatomical outcomes helps to further a mechanistic understanding of Ras/MAPK’s impact on neurodevelopmental outcomes. Further refinement of predictive modeling to estimate individualized child outcomes will advance a precision medicine approach to treating NS, other neurogenetic syndromes, and neuropsychiatric disorders more broadly.

## Introduction

The precision medicine model seeks to transform clinical treatment by combining an individual’s unique environmental, genetic/genomic, clinical, and biomarker data to enhance personalized care[1, 2]. Precision medicine approaches have been applied to treat cancer [3] and cardiovascular disease [4] but applications to psychiatric disorders are lagging. The global disease burden of mental disorders in children and adolescents has increased by 14.9% in recent years bringing the total to 21.5 million disability-adjusted life-years (or, years of life lived with the disability) [5]. Despite this dramatic increase, the field of child psychiatry has been relatively stagnant in terms of developing precision medicine approaches [6]. There are currently no evidence-based precision approaches for attention deficit hyperactivity disorder (ADHD) [7], autism spectrum disorders (ASD) [8] or mood disorders [6]. Here we present a familial modeling framework applied to children with a neurogenetic syndrome affecting the Ras/mitogen-activated protein kinase (Ras/MAPK) as a model for developing precision medicine approaches in childhood neuropsychiatric disorders.

The majority of children with pathogenic variants along the Ras/MAPK pathway develop neuropsychiatric disorders as compared with 15% of children in the general population [9]. Noonan syndrome (NS, 1:2000), the most common genetic disorder affecting the Ras/MAPK pathway, [10] is associated with significantly increased risk for neuropsychiatric disorders such as ADHD (22-48%)[11–13] oppositional defiant disorder (ODD, 19%)[13] and 6-23% have learning disabilities[14]. Children with NS are also at greater risk for higher levels of externalizing behaviors[15], anxiety, and depression[16] relative to peers. However, individual neuropsychiatric profiles vary widely[13], and clinicians and researchers are not yet able to predict who will develop a neurodevelopmental disorder or estimate disease severity. Consequently, current interventions for children with NS are focused on symptomatic treatment, as is the case for most idiopathic disorders, like ADHD in which stimulants are the current standard of treatment[17].

Evidence from our lab and others supports the use of NS as a human model to advance understanding of neuropsychiatric disorders. NS is caused by autosomal dominant mutations of high penetrance in specific genes. Over ∼65% of individuals with NS present with a missence mutation of the *PTPN11* or *SOS1* gene[18] which, in both cases, leads to downstream upregulation of the Ras/MAPK signaling cascade[18–20]. Gene discovery has also demonstrated that the encoded proteins of varied genes involved with ASD and other neuropsychiatric disorder cluster within the Ras/MAPK signaling pathway suggesting that these disorders are associated with enrichment of Ras/MAPK signaling [21, 22]. Finally, NS significantly impacts brain development resulting in altered brain structure[23–25] and functional connectivity[26].

The present cross-sectional analysis aims to better understand variability in clinical and biomarker data while accounting for environmental and genetic factors among children with NS. To that end, our primary analysis sought to model the ‘family variance,’ a summation of inherited cognitive and neuropsychiatric traits as well as common environmental milieu (Figure 1a). We will utilize parent cognitive and behavioral data to model corresponding traits in affected children. Our hypothesis, based on preliminary data in NS and studies in other neurogenetic syndromes,[27–34] is that parent data will allow us to account for common genetic and environmental factors and thus improve the precision of predictive models over and beyond prediction based on genetic diagnosis alone.

**Figure 1.**
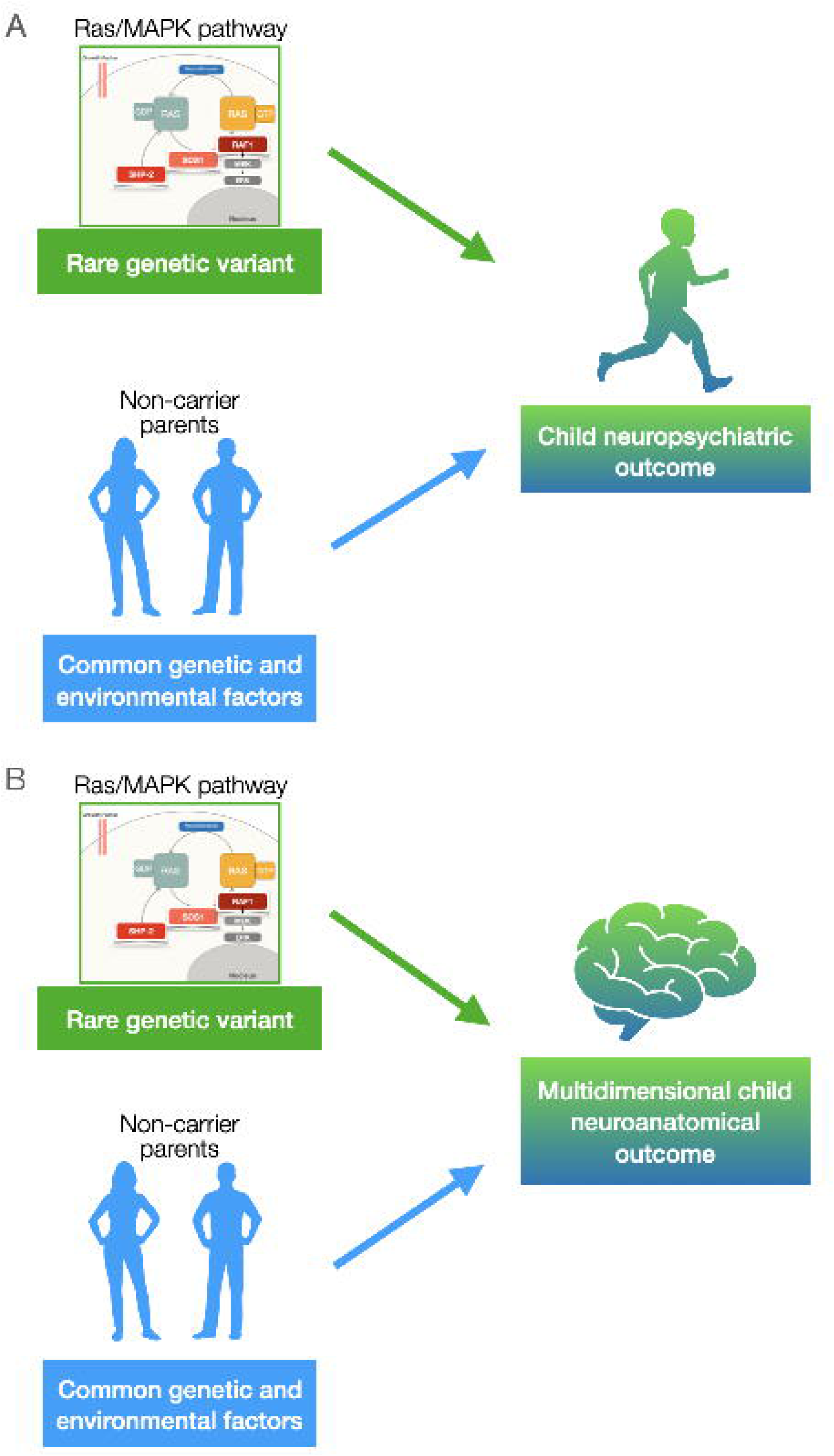
Illustration of the pathogenic variant of large effect and common genetic variance and environmental factors that influence neuropsychiatric outcomes in children with Noonan syndrome. A. Parent traits can be used to predict a child’s behavioral outcome. B. Parent traits can be used to predict a child’s neuroanatomical phenotype which may serve as an intermediate phenotype between pathogenic variants and a child’s cognitive/behavioral outcomes.

Moreover, NS has a large effect on neurobiological development[23, 24, 26], which may represent an important intermediate phenotype (i.e. endophenotype) between Ras/MAPK pathogenic variants and neuropsychiatric phenotypes (Fig 1b.). Yet, because previous research compared children with NS to typically developing peers, we cannot yet disentangle the effects of the pathogenic variant from the effects of common genetic and environmental factors. Thus, our secondary analysis applied machine learning to examine the correspondence between non-carrier parent IQ and multivariate neuroanatomical outcomes in children with NS.

## Materials and Methods

### Participants

We enrolled fifty-three families with children aged 4-12.9 years old (mean = 8.48, SD = 2.12, 34 female) diagnosed with NS and *PTPN11* (N=43) or *SOS1* (N=11) pathogenic variants. Research was performed at the Stanford University School of Medicine and the Institutional Review Board approved all study procedures. Written, informed consent was obtained from a legal guardian for all participants. All participants over 7 years provided assent.

Participants with NS were recruited via the National Noonan Syndrome Foundation, a local network of physicians and advertisements posted on the Stanford University School of Medicine website. TD participants were recruited through two NIH funded studies (HD090209 K23 and HD108684 R01) and a study funded by the Neurofibromatosis Therapeutic Acceleration Program (NTAP) at the Johns Hopkins University School of Medicine using local print media and parent networks. Potential participants across both groups were excluded for premature birth (gestational age <32 weeks), low birth weight (<2000g) or diagnosis of a major psychiatric disorder. Additional exclusion criteria included history of the following: seizures, neurological disorders known to have an impact on cognitive development or brain structure, gross structural brain malformations. All participants were free from MRI contraindications. We estimated power based on previous work demonstrating a correlation between parent and child IQ in XYY syndrome (r=0.63)[34]. Using a more conservative r value of 0.50, the estimated power to detect an association between parent and child IQ for a cohort of 53 families is 0.98. All recruitment and assessments were completed between 2018 and 2022.

### Parent and child assessments

General cognition was assessed for parents and children with age-appropriate versions of the Wechsler Intelligence Scales[35–37]. We utilized the full-scale intelligence quotient (FSIQ) and Block Design, Vocabulary, Matrix Reasoning and Similarities subtest scores. Behavior was assessed via the Achenbach System of Empirically Based Assessment (ASEBA)[38, 39]. We collected behavioral data from one or both parents via the Adult Self Report and, when data from both parents was available, we averaged to create a composite score. For child assessment we collected parent report forms from both parents when possible and scores were averaged to create a composite. For further analysis we used the Diagnostic and Statistical Manual-5th edition (DSM-5) scales that correspond between adult (parent) and child forms: Depressive Problems, Anxiety Problems, Somatic Problems, Attention Deficit/Hyperactivity Problems[40].

### Neuroanatomical variation (child)

All child participants completed behavioral training in a mock MRI scanner to reduce sensitivity to the MRI environment and to reduce motion during the actual scan[41]. Next, child participants completed one of two different (but analogous) T1-weighted anatomical imaging protocols. The first was completed on a GE Healthcare Discovery 3.0 Tesla whole-body MR system using a standard 8-channel head coil (GE Medical Systems, Wilwaukee, WI). Sagittal T1 images were acquired (repetition time 8.2 ms; echo time 3.2 ms; flip angle 12°; field of view 256 × 256 mm; matrix 256 × 256; 176 slices; voxel size = 1.0 × 1.0 × 1.0 mm). The second was performed on a GE 3T SIGNA Premier whole-body MR system using a standard 48-channel head coil (GE Medical Systems, Milwaukee, WI). A 3D magnetization-prepared rapid gradient echo pulse sequence was used to acquire axial T1 images of the brain (repetition time□=□1985 ms, echo time□=□2.8 ms, inversion time□=□900 ms, flip angle□=□8°, slice thickness□=□1.2 mm, field of view = 24.0 cm, acquisition matrix□=□240 × 240, voxel size□=□1.2 × 1.0 × 1.0 mm, and duration□=□4:22.).

FreeSurfer (http://surfer.nmr.mgh.harvard.edu/) (version 5.3 for for the first and 7.1 for the second protocol) was used to delineate 86 gray matter regions (68 cortical, 16 subcortical, 2 cerebellum, See supplement), and compute measures of regional thickness/volume. FreeSurfer is a surface based segmentation pipeline that preserves anatomical variation at the individual level and provides reliable segmentation of cortical, subcortical and cerebellar structures.[42] For the first protocol, bias field correction methods in SPM8 (http://www.fil.ion.ucl. ac.uk/spm) were utilized on the anatomical MRI images prior to processing through the FreeSurfer pipeline. FreeSurfer derived surfaces were examined and adjusted by editors with inter-rater reliability ≥ 0.95 (http://surfer.nmr.mgh.harvard.edu/fswiki/FsTutorial) until the surfaces satisfactorily delineated both the gray/white boundary and pial surface. Image quality requirements included lack of artifacts induced by subject motion, blood flow, or wraparound. Three scans from the first sequence and 0 scans from the second sequence were unusable due to such artifacts.

Freesurfer cortical thickness and subcortical/cerebellar volume metrics were used as data elements in the multivariate modeling.[43, 44] Cortical thickness metrics may represent a superior endophenotype for neurogenetic syndromes when compared to cortical volume[45]. Thus, all cortical structures were summarized according to thickness. Subcortical and cerebellar structures were summarized according to volume, the only available metric in the FreeSurfer parcellation.

Combat harmonization was performed to reduce differences related to technical factors (different scan sequences) while preserving biological variability [46]. The combat method removes non biological variance while increasing the power and reproducibility of statistical analyses in neuroimaging datasets. We performed the harmonization using age, sex and pathogenic variants as biological variables and scan sequence (first or second) as the non-biological, technical variable to be harmonized across. The harmonization procedure was performed on both the parcellated data and the vertex-wise data.

Regional measures were normalized to remove effects of overall brain size, sex, age and pathogenic variant using residuals from linear regression. Due to the different scale for cortical thickness (∼2–4 mm) and subcortical/cerebellar volumes (several hundred mm3) additional normalization included dividing the value for each region by the absolute value of the maximum residual for that region.[43, 44] The resulting value were between −1 and 1 for each of 86 regions.

### Univariate analysis for parent/child outcome correspondence

For each trait we used effect sizes to assess offsets (parent - child scores). Slopes were used to estimate the familial effect on each outcome. For each outcome a regression model was run after centering children and family scores at the mean family score. Thus, model intercepts estimate the average score offset of children relative to parents for each trait. The slopes (B1 coefficient) were compared to 1 (equivalent to a 1- to-1 correspondence between child and family scores and a stable offset across the range of parent scores) and 0 (lack of significant relationship between child and family scores).

Stepwise regression was used to examine the relationship between parent and child traits. Parent trait was the predictor, control variables 1) pathogenic variant (*SOS1* or *PTPN11*) and 2) child age were entered on the first step and the child trait was entered on the final step (outcome measure). Then we performed a second set of regressions for the behavioral traits in which pathogenic variant and age were entered in the first step, FSIQ was entered next, and the child trait was entered in the last step. We performed a final set of regressions in which additional control variables were entered in the first step. The additional variables included child’s sex, child’s birth weight, and child’s cardiac problems (which were added in addition to pathogenic variant and child’s age). The child’s outcome was entered on the final block. In each regression we examined R^2^ change which estimates the percentage of unique variance in child outcome that is accounted for by the corresponding parent measure after accounting for variance in the relevant control variables entered in each regression.

### Multivariate regression analysis

Linear support vector regression[47] (SVR) was used to assess the relationship between parent IQ and child multivariate neuroanatomical outcomes. SVR is a flexible technique that assesses the association among data elements based on linear separation in multidimensional space and can accommodate any type of high-dimensional data. Data elements included metrics from 86 Freesurfer-derived brain regions (See supplement). A set of linearly uncorrelated components was extracted from these elements via principal component analysis. All principal components were input as features in the subsequent SVR. A leave-one-out cross validation was performed to test prediction accuracy. Recursive feature elimination (RFE) was used to reduce the number of features and avoid overfitting. During RFE, the bottom 30% of features, based on the absolute value of their weights, were excluded in an iterative process until model performance started degrading. The RFE procedure was done within each cross-validation set (i.e., nested RFE) to avoid bias in classification accuracy. The regression coefficient (r value) was computed, and significance was tested based on null models using permutation testing (n=1000). Then model performance was assessed using Spearman’s correlation between predicted and observed scores. Finally, the weights of each region’s contribution to the SVR algorithm were examined to identify regional contribution to the overall model.

### Validation procedures

To validate the harmonization across scanning protocols (combat) we performed an analysis using only the “first” scan sequence (N=40) and freesurfer ROIs. We also performed a second validation analysis using freesurfer vertices as data elements (N=327,684 vertices) to test whether the predictive models between parent traits and child’s neuroanatomical variation hold when a much more fine-grained brain parcellation is used.

## Results

Parent/child IQ scores were available from 52 families. Only one parent completed IQ testing in each family (N mothers = 49, N fathers = 4, N missing = 1). Parent/child behavioral scores were available from 51 families: N=45 with both parents providing data and N=6 with one parent providing data.

### Univariate results

Offsets indicated that child cognition (FSIQ scores) was shifted down relative to parent scores: mean difference = 12.535, p<0.001, Cohen’s d= 1.04. For behavior, child scores were shifted up (indicating greater levels of behavior problems) relative to parent scores for depression (d = -0.82), anxiety (d = -0.82), and ADHD problems (d = -0.82, all p’s <0.001, survive Bonferroni correction) but not somatic problems p>0.10, d = -0.57).

Regression results indicate that parent FSIQ was a significant predictor of child IQ after controlling for child pathogenic variant (*PTPN11* or *SOS1*) (p =0.030) Table 1 and Figure 2. Upon examination of verbal and performance subtest data, the model and change in R^2^ were significant only for block design (p =0.002).

**Figure 2.**
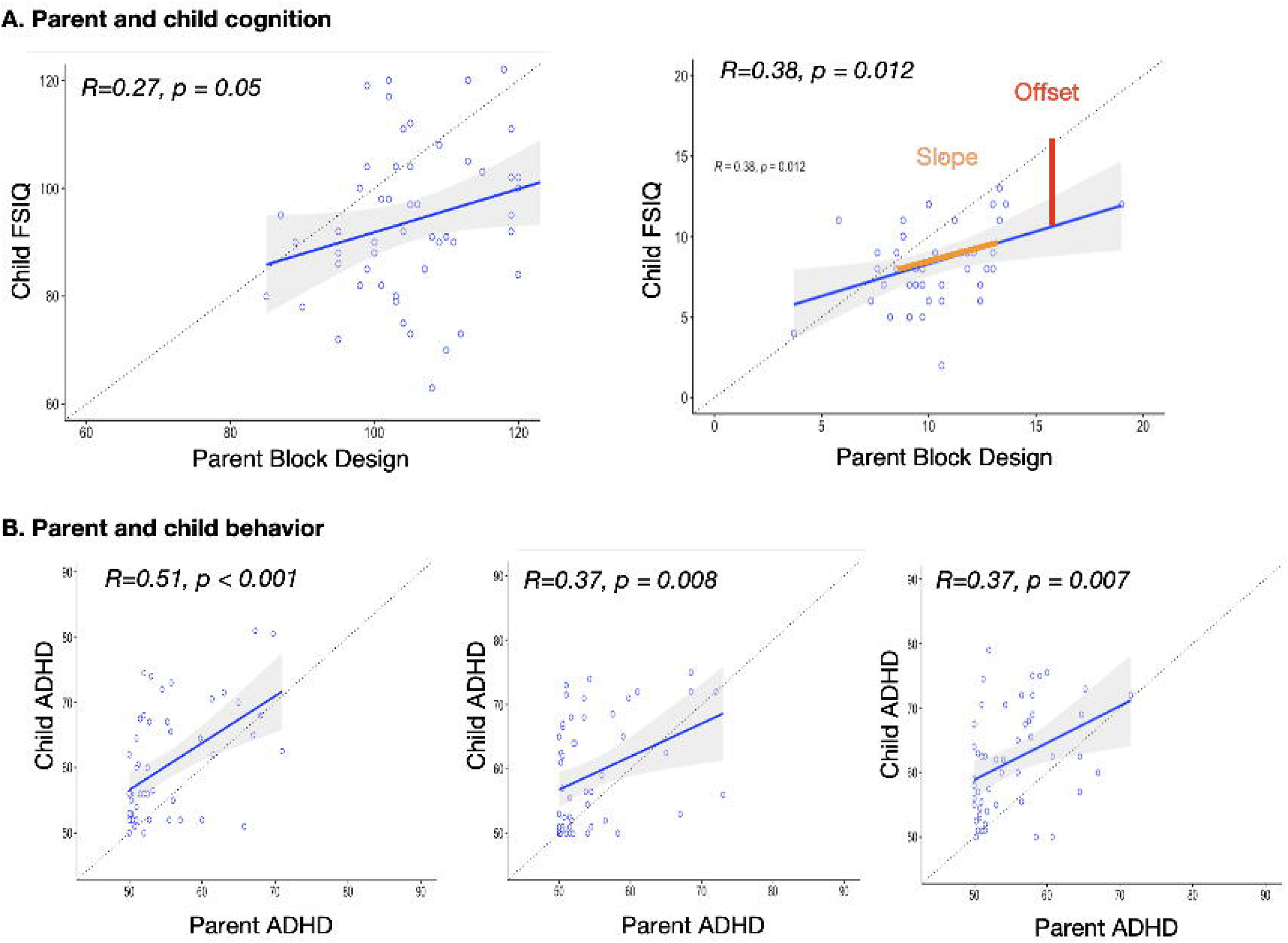
Univariate results A. Linear regression plots for prediction of child cognition from parent cognition. The dashed line represents a hypothetical “identity” line, where one unit change in parent measure = 1 unit change in child measure. The red arrow (shown in the FSIQ plot only, for example) indicates offset, which estimates the familial effect. The solid blue line represents the best-fit regression line. Slope tests for shifting familial effect with parent score, dots represent individual data points for parent and corresponding child measure. Simple correlation and corresponding p values are presented. See below for regression results. Regression framework adapted from Wilson et al. 2021. FSIQ = Wechsler full scale intelligence quotient. Standard scores are presented. For Wechsler subtests, scaled scores are presented.

**Table 1.**
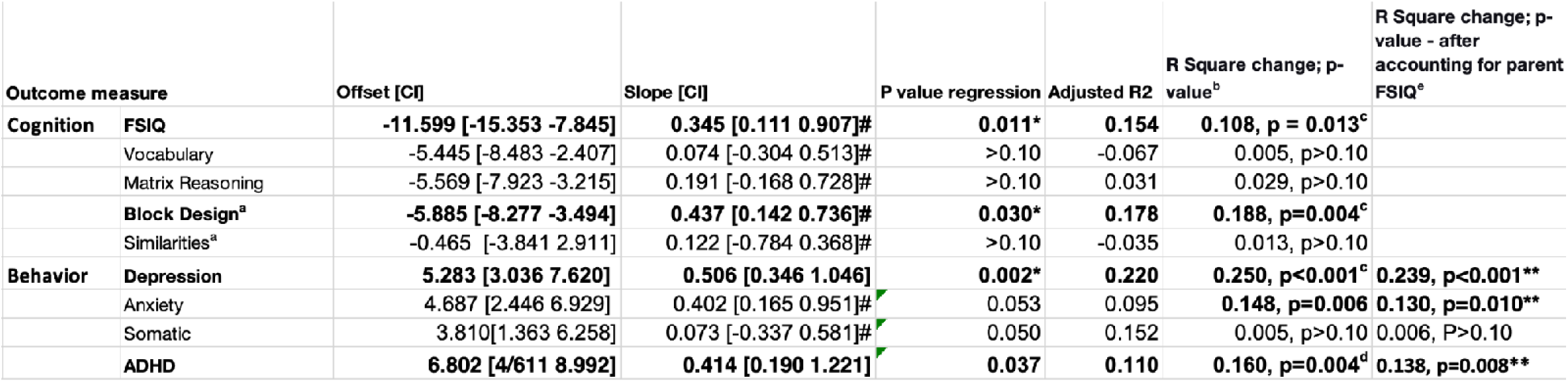
Results of hierarchical linear regression. Covariates child age and pathogenic variant entered in the first block, child outcome in the second block. Cognitive measures include Wechsler scales for full scale IQ (FSIQ) and 4 subtests. Behavior measures are scales from the Achenbach system. CI = 95% confidence interval; *regression model is significant; ** regression model including age, genetic mutation, and parent FSIQ is significant; ^#^Slope different from 1. ^a^N=45, all others are N=53; ^b^Same as the p value for slope in this case. For Behavioral measures, we also tested whether the r squared change value was significant after accounting for the parent FSIQ; ^c^Regression and R^2^ change values are significant with additional covariates: child’s sex, birthweight, cardiac problems. ^d^Regression with additional covariates p = 0.052, r square change p = 0.006. ^e^Due to missing data N=41 for IQ and N=38 for behavior.

| Outcome measure |  | Offset [CI] | Slope [CI] | P value regression | Adjusted R2 | R Square change; p-value<sup>b</sup> | R Square change; p-value - after accounting for parent FSIQ<sup>e</sup> |
| --- | --- | --- | --- | --- | --- | --- | --- |
| Cognition | FSIQ | **-11.599 [-15.353 -7.845]** | **0.345 [0.111 0.907]#** | **0.011\*** | **0.154** | **0.108, p = 0.013<sup>c</sup>** |  |
|  | Vocabulary | -5.445 [-8.483 -2.407] | 0.074 [-0.304 0.513]# | >0.10 | -0.067 | 0.005, p>0.10 |  |
|  | Matrix Reasoning | -5.569 [-7.923 -3.215] | 0.191 [-0.168 0.728]# | >0.10 | 0.031 | 0.029, p>0.10 |  |
|  | **Block Design<sup>a</sup>** | **-5.885 [-8.277 -3.494]** | **0.437 [0.142 0.736]#** | **0.030\*** | **0.178** | **0.188, p=0.004<sup>c</sup>** |  |
|  | Similarities<sup>a</sup> | -0.465 [-3.841 2.911] | 0.122 [-0.784 0.368]# | >0.10 | -0.035 | 0.013, p>0.10 |  |
| Behavior | **Depression** | **5.283 [3.036 7.620]** | **0.506 [0.346 1.046]** | **0.002\*** | **0.220** | **0.250, p<0.001<sup>c</sup>** | **0.239, p<0.001\*\*** |
|  | Anxiety | 4.687 [2.446 6.929] | 0.402 [0.165 0.951]# | 0.053 | 0.095 | **0.148, p=0.006** | **0.130, p=0.010\*\*** |
|  | Somatic | 3.810[1.363 6.258] | 0.073 [-0.337 0.581]# | 0.050 | 0.152 | 0.005, p>0.10 | 0.006, P>0.10 |
|  | **ADHD** | **6.802 [4/611 8.992]** | **0.414 [0.190 1.221]** | **0.037** | **0.110** | **0.160, p=0.004<sup>d</sup>** | **0.138, p=0.008\*\*** |

Subsequent regression results indicated that parent behavior was a significant predictor of child behavior after controlling for child pathogenic variant (*PTPN11* or *SOS1*) and parent FSIQ. Results were significant for depression (p<0.001), anxiety (p =0.008) and ADHD problems (p =0.013). Due to missing covariate data, analysis with additional covariates (child’s sex, birth weight, and cardiac problems) included 41 families for IQ and 38 for behavior. As indicated in Table 1, models for FSIQ, Block design and Depression remained significant after accounting for the additional variables. R^2^ change was significant for FSIQ, Depression, Anxiety and ADHD. *Multivariate results*

Parent FSIQ was not significantly associated with total brain volume or with mean cortical thickness after accounting for child age, sex, and pathogenic variant (p’s >0.10). The SVR analysis indicated that parent IQ was significantly associated with child neuroanatomical variation using a leave one out approach. RFE indicated a model with one feature (one principal component) was optimal (R=0.34, p=0.0015; r_s_ =0.403, p=0.004). Regions with the highest contribution to the classification accuracy (arbitrary threshold for weight = 0.5) and visualized in Figure 3 and the weights of each region are listed in Table 2. The SVR models are based on the pattern of variation in neuroanatomy; they can not specify directionality of associations with individual brain regions. Therefore, we performed post-hoc correlations between the aforementioned regions with weights >0.5 and parent IQ (Supplementary Table 3). Validation analysis with vertex-based data, and with only data from the first anatomical imaging protocol revealed significant associations between parent IQ and child neuroanatomical variation. Using vertex-based parcellation a model with 5 features was optimal (R=0.34, p=0.011; r_s_ =0.36, p=0.01). Using only data from the first imaging protocol (N=40) a model with two features was optimal (R=0.36, p=0.008, r_s_ =0.35, p=0.026).

**Figure 3.**
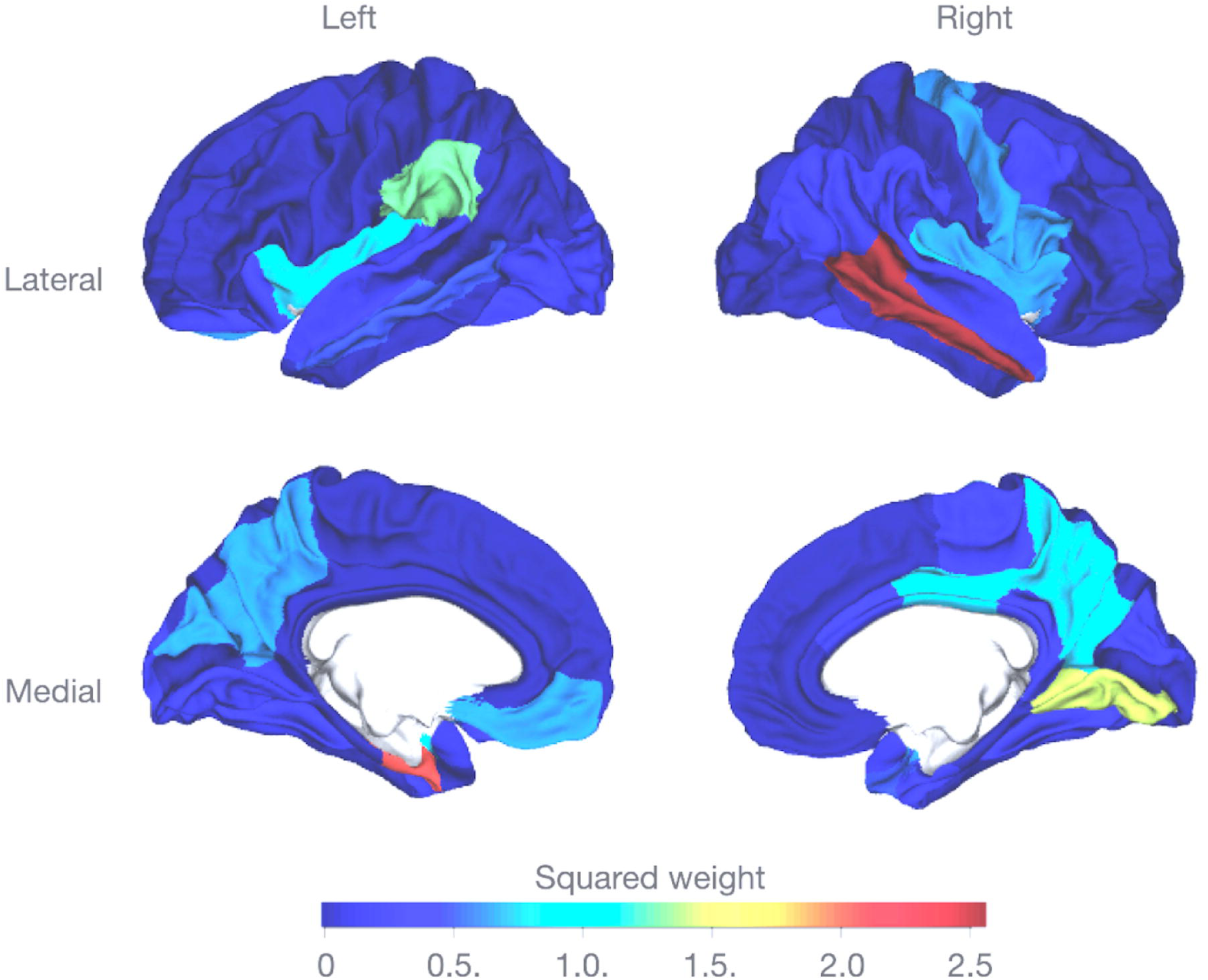
Multivariate results. Parent’s IQ was significantly associated with child neuroanatomical variation. Each Freesurfer region of interest is colored according to the squared weights representing each region’s contribution to the optimal model.

**Table 2.**
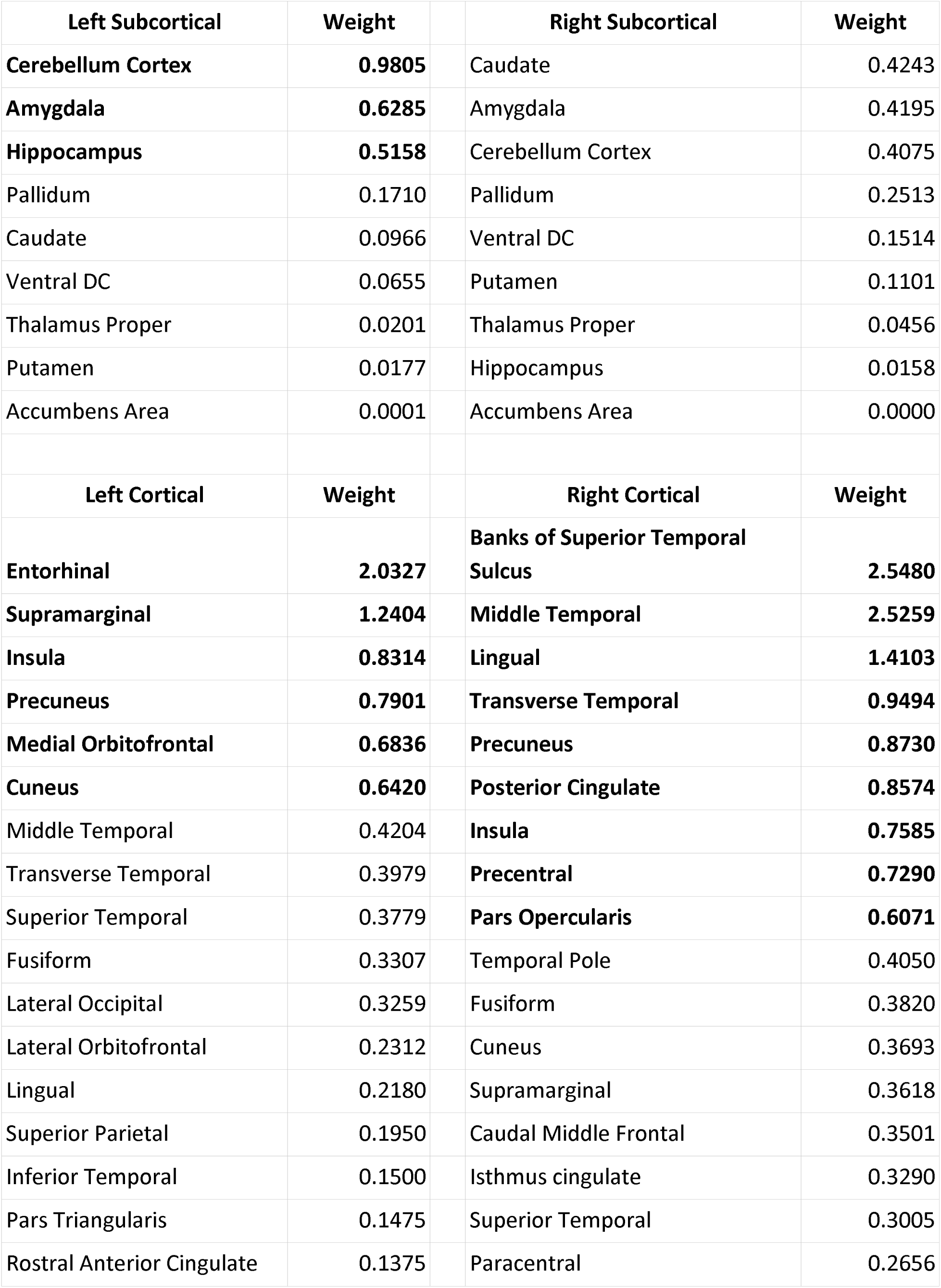
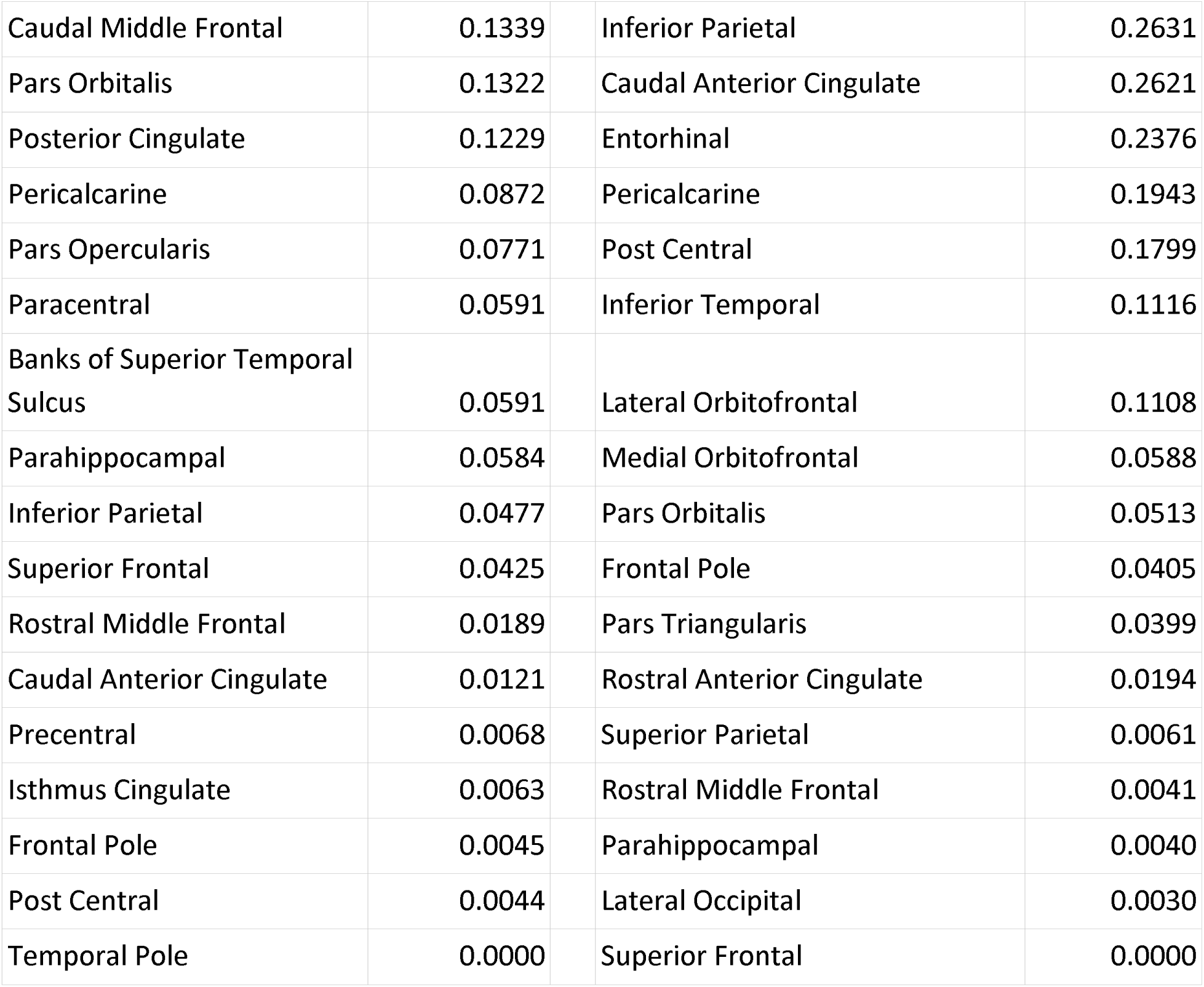
Weights for each region in the optimal SVR model. Regions with weight above threshold 0.50 are bolded. Regions are sorted first according to cortical/subcortical location then according to their weights in the optimal model.

## Discussion

Here we present original research demonstrating that parent traits are useful in understanding child neuropsychiatric and neuroanatomical outcomes. We address two critical aspects of precision medicine - clinical presentation (neuropsychiatric symptoms) and biomarker data (MRI-based neuroanatomical outcomes) using NS as a case study and human model system. Using univariate predictive modeling, we demonstrate that parents’ cognition, depression, anxiety, and ADHD symptoms are significantly associated with the levels of those symptoms in children with NS. Using multivariate machine learning, we demonstrate that parent cognition was significantly associated with the multivariate pattern of anatomical development in children with NS and underscore the importance of amygdala and temporal lobe regions.

Utilizing parent traits in a predictive framework affords control for familial factors and thus provides a more individualized estimate of expected child outcomes. This framework has been used before in XYY syndrome[34]. Measuring parent traits for prediction is a cost effective, noninvasive method that may help clinicians pinpoint child outcomes. With this framework, we provide novel information above and beyond what can be concluded from case/control studies.

Our analysis revealed several important themes. First, we demonstrated a variable familial effect in children with NS across neuropsychiatric symptoms with the greatest effect in cognition (full scale IQ), ADHD, and depression. While weaknesses in cognition and ADHD are well documented for children with NS[13, 48, 49], our results suggest a familial contribution to the child’s behavioral phenotype, above and beyond what is accounted by the genetic variant of large effect (*PTPN11*/*SOS1*). Mood and anxiety problems seem to emerge in older children/adolescents with NS[16], yet these neuropsychiatric symptoms are greatly understudied in this population. Given the high heritability of mood and anxiety disorders in the general population[50], it is important to understand how parent symptoms are associated with child symptoms in NS. Together, our modeling framework provides a comprehensive estimation of the effect of genetic mutation in the context of background genetic and environmental factors.

Our univariate regression analysis demonstrated that parent IQ (especially visuospatial/motor abilities), mood, anxiety, and ADHD are related to child outcomes in these domains. The associations with mood, anxiety, and ADHD were also significant after accounting for parent full scale IQ suggesting that a parent’s IQ was not driving the differences in these symptoms. We also demonstrate that the associations with mood and ADHD are significant after accounting for additional confounders, including child’s sex, birthweight, and cardiac problems. Associations with anxiety reached trend-level (p=0.052) and the change in R^2^ remained significant with these additional covariates. Together, these results suggest that a parent’s traits are useful for predicting not only a child’s cognition but their behavioral symptoms as well. Additionally, for cognitive traits, we found evidence for shifting familial effect - the gap between a parent and child’s IQ became greater when the parent’s IQ was higher (Figure 2). This suggests a potential ceiling in terms of the level of cognitive skill a child with NS is able to attain. In other words, the child’s IQ score is not able to keep up with what would be predicted based on the parent’s IQ alone.

Understanding how parent traits influence neuroanatomical outcomes helps to further a mechanistic understanding of Ras/MAPK’s impact on neurodevelopmental outcomes. We highlighted several regions that were particularly strongly weighted in the model: bilateral temporal and occipital regions and left hemisphere subcortical/cerebellar regions. Our previous study has indicated that children with NS, relative to their typically developing peers, demonstrated altered patterns of altered morphology for several of these regions: left entorhinal cortex, left supramarginal gyrus, the cuneus and precuneus as well as right hemisphere lingual, posterior cingulate precentral gyri and right insula.[51] These regions may be uniquely influenced by both NS pathogenic variants and parent IQ. Our post-hoc univariate correlations did not reveal a significant association between parent IQ and thickness of these regions, supporting the hypothesis that these regions are more prone to the effects of the genetic variant (*PTPN11*/*SOS1*). However, it is possible that these analyses do not completely describe the complex interrelationship between a child’s pathogenic variant, parent IQ, and child’s neuroanatomical outcome. It is also worth reiterating that the SVR model tests for associations between parent IQ and the multivariate pattern of child neuroanatomy. Thus, it is not designed to isolate specific regions. In fact, the association between a parent’s IQ and a child’s multivariate pattern of neuroanatomy suggests a powerful potential predictive relationship with impacts on global brain development.

On the other hand, the left amygdala, right bank of the superior temporal sulcus and the right middle temporal gyrus were associated with parent cognition in our SVR but did not show significantly altered anatomy in the previous study.[51] Further, thickness of these regions correlated significantly and positively with parent IQ in our post hoc analysis (with and without controlling for child’s pathogenic variant). These findings provide evidence that these regions are influenced by parent cognition but not NS pathogenic variants. The amygdala, via connections with the frontal cortex plays a major role in social cognitive processes, including social learning and decision making[52]. The left superior temporal sulcus subserves linguistic, cognitive and social functions including processing faces and auditory stimuli[53–55]. The lingual gyrus subserves functions related to vision but it also plays a role in inhibition[56, 57]. Both social cognition and inhibition are areas of significant weakness for children with NS[13, 58]. Our findings indicate that parents with higher IQ scores may be able to provide a more enriched environment that supports social cognitive and inhibition development, thus influencing their child’s superior temporal sulcus and lingual gyrus anatomical development. These results might also indicate that educational interventions can influence a child’s social cognition and inhibition developmental trajectories. Due to the cross-sectional nature of the present work and past literature we cannot speak to the directionality of skill compared to anatomical development (likely there is a reciprocal process by which neuroanatomy and cognitive skills develop). These are nonetheless interesting preliminary insights which can be further tested with longitudinal research.

Demonstrating that familial modeling can be useful for predicting outcomes in NS is important for several reasons. First, it can help us better understand pathways to the neuropsychiatric and neuroanatomical phenotype specifically in children with NS. Second, we know that prediction of cognition/behavior using parent scores generalizes to other neurogenetic syndromes including XYY[34] and fragile X syndrome[28]. Our work and previous work indicates that these models may generalize to other relatively common RASopathies such as neurofibromatosis 1 (NF1; 1:3000). The association between a parent’s IQ and child’s neuroanatomical outcome has not been tested before. This initial ‘proof of concept’ in NS indicates that these models are significant and robust. We performed two different validations: 1) with a different (finer) anatomical parcellation, 2) with a subset of participant data from a single scan protocol. Both validations indicated similar model performance and the utility of a parent’s IQ in predicting a child’s neuroanatomical variation. Third, we demonstrated the ability to predict complex, multidimensional anatomical development in children using a less invasive, cost-effective measure of parent IQ.

Our study included a relatively small sample size for predictive analysis and machine learning but a relatively large sample for rare neurogenetic syndromes such as NS. Future work in other NS populations will be important. Further, we did not examine parent or child genetics beyond the NS pathogenic variant. Knowledge of both parent and child genetics will be important for further refinement of predictive modeling. Finally, we did not measure parent neuroanatomy which may also hold mechanistic clues to a child’s brain development as previously demonstrated for depression[59] and schizophrenia[60].

Utilizing parent traits in a predictive framework affords control for familial factors and thus provides a more individualized estimate of expected child outcomes. Understanding how parent traits influence a child’s neuroanatomy may help to better understand a child’s neuroanatomical development. This is because the brain outcomes can be considered an intermediate phenotype between genetic factors and neurodevelopmental outcomes. In sum, our work helps to further a mechanistic understanding of Ras-MAPK’s impact on neuropsychiatric and neuroanatomical outcomes. Further refinement of predictive modeling to estimate individualized outcomes will advance a precision medicine approach to treating NS, other neurogenetic syndromes, and neuropsychiatric disorders more broadly.

## Supporting information

Suppemental

## Data Availability

The final dataset will be stripped of all identifiers and made available to qualified investigators upon request.

## Acknowledgements

This work was supported by the National Institute of Child Health and Human Development (#HD090209 K23 and #HD108684 R01). Tamar Green was also supported by the Stephen Bechtel Endowed Faculty Scholar in Pediatric Translational Medicine, Stanford Maternal & Child Health Research Institute and by funding from the Neurofibromatosis Therapeutic Acceleration Program (NTAP) at the Johns Hopkins University School of Medicine. Its contents are solely the responsibility of the authors and do not necessarily represent the official views of The Johns Hopkins University School of Medicine.

