## Supplementary material for "A familial modeling framework for advancing precision medicine in neuropsychiatric disorders: A study in children with RASopathies": Suppemental

Supplemental Methods

*Recruitment and exclusion criteria*

Participants with NS were recruited via the National Noonan Syndrome Foundation, a local network of physicians and advertisements posted on the Stanford University School of Medicine website. TD participants were recruited through two NIH funded studies (HD090209 K23 and HD108684 R01) and a study funded by the Neurofibromatosis Therapeutic Acceleration Program (NTAP) at the Johns Hopkins University School of Medicine using local print media and parent networks. Potential participants across both groups were excluded for premature birth (gestational age <32 weeks), low birth weight (<2000g) or diagnosis of a major psychiatric disorder. Additional exclusion criteria included history of the following: seizures, neurological disorders known to have an impact on cognitive development or brain structure, gross structural brain malformations. All participants were free from MRI contraindications. We estimated power based on previous work demonstrating a correlation between parent and child IQ in XYY syndrome (r=0.63)[34]. Using a more conservative r value of 0.50, the estimated power to detect an association between parent and child IQ for a cohort of 53 families is 0.98. All recruitment and assessments were completed between 2018 and 2022.

*Anatomical MRI data collection and preprocessing (child)*

Child participants completed one of two different (but analogous) T1-weighted anatomical imaging protocols. The first was completed on a GE Healthcare Discovery 3.0 Tesla whole-body MR system using a standard 8-channel head coil (GE Medical

Supplementary Table 1 Freesurfer brain regions

| Region name | Type |
|---|---|
| Cerebellum cortex | subcortical volume |
| Thalamus | subcortical volume |
| Caudate | subcortical volume |
| Putamen | subcortical volume |
| Pallidum | subcortical volume |
| Hippocampus | subcortical volume |
| Amygdala | subcortical volume |
| Accumbens area | subcortical volume |
| Ventral diencephalon | subcortical volume |
| Superior parietal | cortical thickness |
| Caudal anterior cingulate | cortical thickness |
| Cuneus | cortical thickness |
| Parahippocampal | cortical thickness |
| Isthmus cingulate | cortical thickness |
| Rostral Anterior Cingulate | cortical thickness |
| Pericalcarine | cortical thickness |
| Transverse temporal | cortical thickness |
| Caudal middle frontal | cortical thickness |
| Fusiform | cortical thickness |
| Pars triangularis | cortical thickness |
| Temporal pole | cortical thickness |
| Post central | cortical thickness |
| Superior temporal | cortical thickness |
| Middle temporal | cortical thickness |
| Rostral middle frontal | cortical thickness |
| Pars orbitalis | cortical thickness |
| Inferior temporal | cortical thickness |
| Frontal pole | cortical thickness |
| Posterior cingulate | cortical thickness |
| Medial orbitofrontal | cortical thickness |

| | |
|---|---|
| Lateral occipital | cortical thickness |
| Bank of the superior temporal suclus | cortical thickness |
| Paracentral | cortical thickness |
| Insula | cortical thickness |
| Precentral | cortical thickness |
| Supramarginal | cortical thickness |
| Lingual | cortical thickness |
| Pars opercularis | cortical thickness |
| Inferiorparietal | cortical thickness |
| Entorhinal | cortical thickness |
| Superior frontal | cortical thickness |
| Lateral orbitofrontal | cortical thickness |
| Precuneus | cortical thickness |

Supplementary Table 2.

| Left Subcortical | Weight | | Right Subcortical | Weight |
|---|---|---|---|---|
| **Cerebellum Cortex** | **0.9805** | | Caudate | 0.4243 |
| **Amygdala** | **0.6285** | | Amygdala | 0.4195 |
| **Hippocampus** | **0.5158** | | Cerebellum Cortex | 0.4075 |
| Pallidum | 0.1710 | | Pallidum | 0.2513 |
| Caudate | 0.0966 | | Ventral DC | 0.1514 |
| Ventral DC | 0.0655 | | Putamen | 0.1101 |
| Thalamus Proper | 0.0201 | | Thalamus Proper | 0.0456 |
| Putamen | 0.0177 | | Hippocampus | 0.0158 |
| Accumbens Area | 0.0001 | | Accumbens Area | 0.0000 |
| **Left Cortical** | **Weight** | | **Right Cortical** | **Weight** |
| **Entorhinal** | **2.0327** | | **Banks of Superior Temporal Sulcus** | **2.5480** |
| **Supramarginal** | **1.2404** | | **Middle Temporal** | **2.5259** |
| **Insula** | **0.8314** | | **Lingual** | **1.4103** |
| **Precuneus** | **0.7901** | | **Transverse Temporal** | **0.9494** |
| **Medial Orbitofrontal** | **0.6836** | | **Precuneus** | **0.8730** |
| **Cuneus** | **0.6420** | | **Posterior Cingulate** | **0.8574** |
| Middle Temporal | 0.4204 | | **Insula** | **0.7585** |
| Transverse Temporal | 0.3979 | | **Precentral** | **0.7290** |
| Superior Temporal | 0.3779 | | **Pars Opercularis** | **0.6071** |
| Fusiform | 0.3307 | | Temporal Pole | 0.4050 |
| Lateral Occipital | 0.3259 | | Fusiform | 0.3820 |
| Lateral Orbitofrontal | 0.2312 | | Cuneus | 0.3693 |
| Lingual | 0.2180 | | Supramarginal | 0.3618 |
| Superior Parietal | 0.1950 | | Caudal Middle Frontal | 0.3501 |
| Inferior Temporal | 0.1500 | | Isthmus cingulate | 0.3290 |

| | | | | |
|---|---|---|---|---|
| Pars Triangularis | 0.1475 | | Superior Temporal | 0.3005 |
| Rostral Anterior Cingulate | 0.1375 | | Paracentral | 0.2656 |
| Caudal Middle Frontal | 0.1339 | | Inferior Parietal | 0.2631 |
| Pars Orbitalis | 0.1322 | | Caudal Anterior Cingulate | 0.2621 |
| Posterior Cingulate | 0.1229 | | Entorhinal | 0.2376 |
| Pericalcarine | 0.0872 | | Pericalcarine | 0.1943 |
| Pars Opercularis | 0.0771 | | Post Central | 0.1799 |
| Paracentral | 0.0591 | | Inferior Temporal | 0.1116 |
| Banks of Superior Temporal Sulcus | 0.0591 | | Lateral Orbitofrontal | 0.1108 |
| Parahippocampal | 0.0584 | | Medial Orbitofrontal | 0.0588 |
| Inferior Parietal | 0.0477 | | Pars Orbitalis | 0.0513 |
| Superior Frontal | 0.0425 | | Frontal Pole | 0.0405 |
| Rostral Middle Frontal | 0.0189 | | Pars Triangularis | 0.0399 |
| Caudal Anterior Cingulate | 0.0121 | | Rostral Anterior Cingulate | 0.0194 |
| Precentral | 0.0068 | | Superior Parietal | 0.0061 |
| Isthmus Cingulate | 0.0063 | | Rostral Middle Frontal | 0.0041 |
| Frontal Pole | 0.0045 | | Parahippocampal | 0.0040 |
| Post Central | 0.0044 | | Lateral Occipital | 0.0030 |
| Temporal Pole | 0.0000 | | Superior Frontal | 0.0000 |

Weights for each region in the optimal SVR model. Regions with weight above threshold 0.50 are bolded. Regions are sorted according to their weights.

| Region name | Correlation with parent IQ | Correlation with parent IQ controlling for pathogenic variant | Previous findings: Noonan syndrome vs typically developing |
|---|---|---|---|
| Right Bankssts Thickness | .375** | 0.361* | - |
| Right Middle Temporal Thickness | .552** | 0.498** | - |
| Left Entorhinal Thickness | -0.106 | -0.163 | ↓ volume and surface area |
| Right Lingual Thickness | 0.02 | -0.051 | ↓ volume and cortical thickness |
| Left Supramarginal Thickness | 0.194 | 0.191 | ↑ surface area |
| Left Cerebellum Cortex | 0.25 | 0.262 | - |
| Right Transversetemporal Thickness | 0.229 | 0.211 | - |
| Right Precuneus Thickness | 0.105 | 0.109 | - |
| Right Posteriorcingulate Thickness | 0.106 | 0.088 | ↓ volume |
| Left Insula | 0.009 | 0.0004 | ↑ thickness |
| Left Precuneus Thickness | 0.019 | 0.056 | ↓ volume |
| Right Insula Thickness | -0.111 | -0.129 | ↑ thickness |
| Right Precentral Thickness | 0.049 | 0.089 | ↓ thickness and volume |
| Left Medial Orbitofrontal Thickness | 0.06 | 0.103 | - |
| Left Cuneus Thickness | 0.114 | 0.099 | ↓ volume and surface area |
| Left Amygdala | .301* | 0.317* | - |
| Right Parsopercularis Thickness | -0.05 | -0.036 | - |
| Left Hippocampus | -0.149 | -0.11 | - |

Supplementary Table 3. Post-hoc correlation results and summary of previous findings for each region highlighted in the optimal model for the association between parent IQ and child multivariate neuroanatomical outcome. Regions are listed based on their weights in the model. Correlation with parent IQ: Pearson correlation coefficient values between region metric and parent IQ are presented. Correlation with parent IQ controlling for pathogenic variant: partial correlation between region metric and parent IQ while controlling for child's pathogenic variant (*PTPN11* or *SOS1*). *p<0.05, two tailed; **p<0.01, two-tailed. Previous findings are summarized from[51], if no findings are listed then there was no group difference reported between Noonan syndrome and typically developing children. The Cerebellum was not investigated in the previous study.
